## Supplemental file for "Spatial Clusters of Caesarean Sections across India - Insights from NFHS Data"

### 1. Additional Tables

Table S1: Analysis of Variance (ANOVA) for proportion of c-section births by place of residence, place of birth and NFHS survey

| Parameter | Sum of Squares (SS) | Degrees of freedom (df) | Mean Squares (MS) | F-statistic | p-value | Partial eta squared (ƞ²) | Partial eta squared (ƞ²) CI low | Partial eta squared (ƞ²) CI high |
| --- | --- | --- | --- | --- | --- | --- | --- | --- |
| Place of residence | 2,398.26 | 1.00 | 2,398.26 | 13.86 | 0.000 | 0.05 | 0.02 | 1.00 |
| NFHS | 1,277.46 | 1.00 | 1,277.46 | 7.38 | 0.007 | 0.03 | 0.00 | 1.00 |
| Place of birth | 48,421.13 | 1.00 | 48,421.13 | 279.76 | 0.000 | 0.52 | 0.45 | 1.00 |
| Residuals | 45,000.33 | 260.00 | 173.08 | --- | --- | --- | --- | --- |

The Analysis of Variance (ANOVA) suggests that the main effect of place_of_residence is statistically significant and small (F(1, 260) = 13.86, p < .001; Eta2 (partial) = 0.05, 95% CI [0.02,1.00]).The main effect of NFHS survey was also statistically significant and small (F(1,260) = 7.38, p = 0.007; Eta2 (partial) = 0.03, 95% CI [4.28e-03, 1.00]). The main effect of cleaned_indicator is statistically significant and large (F(1, 260) = 279.76, p < .001; Eta2 (partial) = 0.52, 95% CI [0.45,1.00]). See Table (Table S1)

Table S2: Clustering of C-sections (%) across public and private facilities in India.

| Parameter | Facility | NFHS | G Statistic | Z Score | p-value |
| --- | --- | --- | --- | --- | --- |
| Caesarean Sections | Overall | NFHS-4 | 0.01099 | 18.67214 | <0.0001 |
| Caesarean Sections | Overall | NFHS-5 | 0.01081 | 19.13736 | <0.0001 |
| Caesarean Sections | Public | NFHS-4 | 0.00810 | 8.28497 | <0.0001 |
| Caesarean Sections | Public | NFHS-5 | 0.00808 | 9.00839 | <0.0001 |
| Caesarean Sections | Private | NFHS-4 | 0.01082 | 19.18451 | <0.0001 |
| Caesarean Sections | Private | NFHS-5 | 0.00749 | 17.28355 | <0.0001 |

### 2. Additional Figures

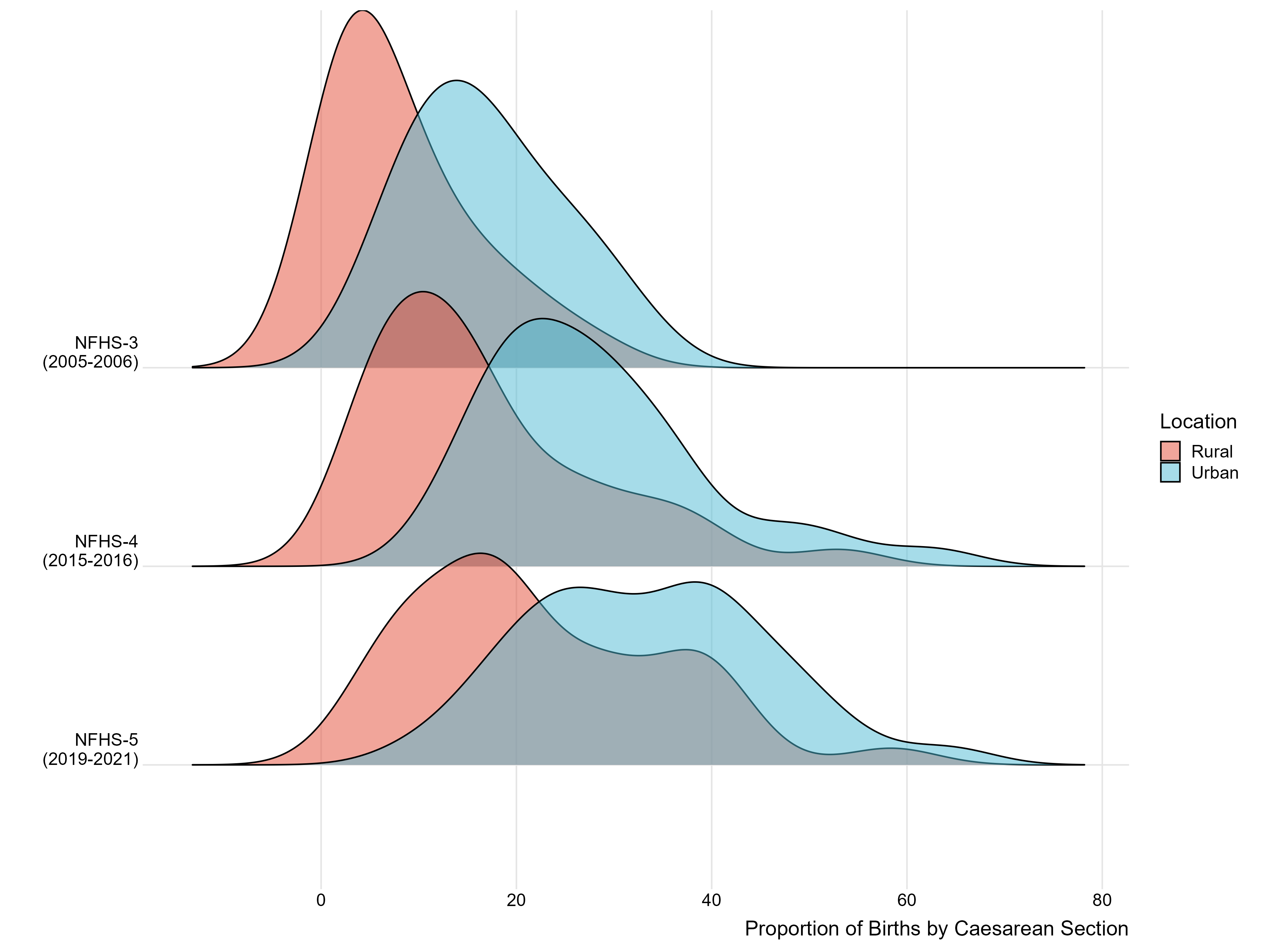

Figure S1: Distribution of the density of the proportion of c-section births across rural and urban India

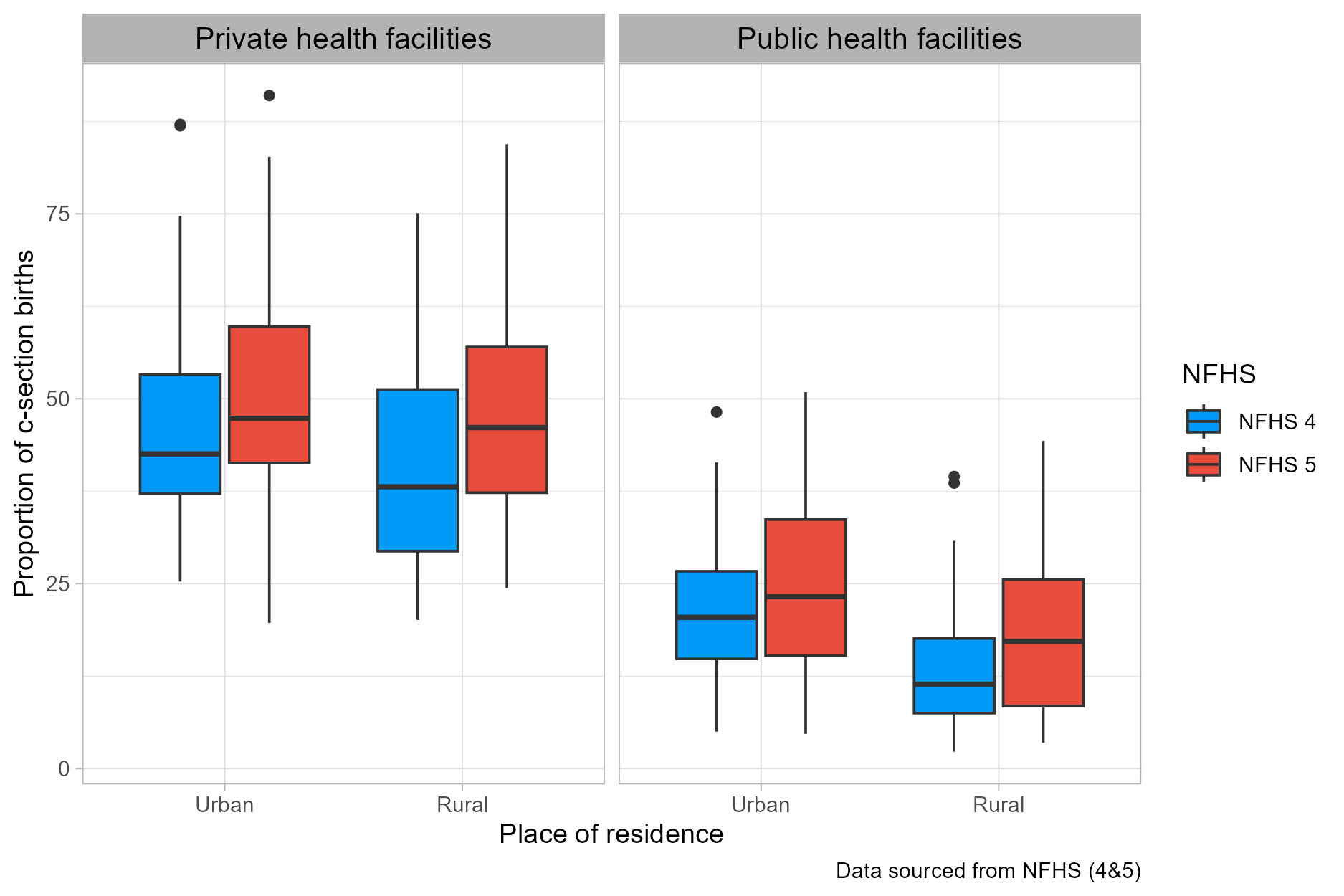

Figure S2: Increase in median proportion of c-section births (%) by place of residence, place of birth and NFHS survey

### 3. Packages Used

The R packages used in the study are as follows:

1. pak
2. here
3. fs
4. rvest
5. tidyverse
6. sf
7. patchwork
8. rgeoda
9. gstat
10. gt
11. gtExtras
12. gtsummary
13. rmapshaper
14. tabulapdf
15. pdftools
16. janitor
